# Genome-wide study of gene-by-sex interactions identifies risks for cleft palate

**DOI:** 10.1101/2024.05.01.24306701

**Authors:** Kelsey Robinson, Randy Parrish, Wasiu Lanre Adeyemo, Terri H. Beaty, Azeez Butali, Carmen J. Buxó, Lord JJ Gowans, Jacqueline T. Hecht, Lina Moreno, Jeffrey C. Murray, Gary M. Shaw, Seth M. Weinberg, Harrison Brand, Mary L. Marazita, David J. Cutler, Michael P. Epstein, Jingjing Yang, Elizabeth J. Leslie

## Abstract

Structural birth defects affect 3-4% of all live births and, depending on the type, tend to manifest in a sex-biased manner. Orofacial clefts (OFCs) are the most common craniofacial structural birth defects and are often divided into cleft lip with or without cleft palate (CL/P) and cleft palate only (CP). Previous studies have found sex-specific risks for CL/P, but these risks have yet to be evaluated in CP. CL/P is more common in males and CP is more frequently observed in females, so we hypothesized there would also be sex-specific differences for CP. Using a trio-based cohort, we performed sex-stratified genome-wide association studies (GWAS) based on proband sex followed by a genome-wide gene-by-sex (GxS) interaction testing. There were 13 loci significant for GxS interactions, with the top finding in *LTBP1* (RR=3.37 [2.04 - 5.56], p=1.93x10^-6^). LTBP1 plays a role in regulating TGF-B bioavailability, and knockdown in both mice and zebrafish lead to craniofacial anomalies. Further, there is evidence for differential expression of *LTBP1* between males and females in both mice and humans. Therefore, we tested the association between the imputed genetically regulated gene expression of genes with significant GxS interactions and the CP phenotype. We found significant association for *LTBP1* in cell cultured fibroblasts in female probands (p=0.0013) but not in males. Taken altogether, we show there are sex-specific risks for CP that are otherwise undetectable in a combined sex cohort, and *LTBP1* is a candidate risk gene, particularly in females.

## Introduction

Structural birth defects occur in 3-4% of all live births (Lary and Paulozzi 2001; Sokal, Tata, and Fleming 2014) and account for 20% of infant mortality (Copeland and Kirby 2007; Ely and Driscoll 2019). There are several factors associated with increased risk for structural birth defects, including maternal and paternal ages, environmental exposures, and genetic variation. In general, more males are born with structural birth defects than females (Tennant et al. 2011; Cui et al. 2005). Although there are varying degrees of both phenotypic and genetic heterogeneity among different structural birth defects, this sex discrepancy suggests inherent sex-specific risks. One such example is that of orofacial clefts (OFCs). For clefts involving the upper lip (cleft lip with or without a cleft palate, CL/P), males are affected at a rate about twice that of females, while for cleft palate (CP), females are affected more frequently (Nasreddine, El Hajj, and Ghassibe-Sabbagh 2021; Mai et al. 2019).

OFCs are a common congenital anomaly, occurring in approximately 1 in 1000 live births (Mai et al. 2019). The need for surgical intervention at an early age, advanced orthodontic care, and speech and hearing therapies contribute to a substantial individual and public health burden. As such, better understanding of their etiology can lead to prevention and/or improved health outcomes. OFCs are etiologically heterogenous, with environmental exposures such as maternal multivitamin use (Yoshida et al. 2020), and maternal alcohol consumption (DeRoo et al. 2016) or smoking (Kummet et al. 2016; Little, Cardy, and Munger 2004) contributing to occurrence risk, as well as genetic factors. CP is highly heritable with estimates around 80-90% (Sivertsen et al. 2008; Grosen et al. 2010), but CP is still understudied compared to CL/P (Dixon et al. 2011; Leslie and Marazita 2013; Carlson et al. 2018; Awotoye et al. 2022).

Several models exist to explain how sex biases manifest in structural birth defects and in CP. An X-linked CP with ankyloglossia phenotype has been reported with variants in *TBX22*, primarily affecting males but appearing to follow a semi-dominant X-linked inheritance pattern (Marçano et al. 2004). Although seemingly counterintuitive, aberrant and incomplete X inactivation secondary to p53 loss has been suggested as a mechanism for increased incidence of neural tube defects in females (Delbridge et al. 2019). Further, there are other X-linked disorders in which female heterozygotes exhibit a wide spectrum of phenotypes as compared to more severely affected males (Pinto et al. 2010; Savige et al. 2016). Another likely contributing factor is the timing of palatal closure in embryogenesis: female embryos undergo this process approximately one week later than males (Burdi and Silvey 1969). This delay in closure allows more time for disruption of the growth and fusion of palatal shelves compared to male embryos and may ultimately contribute to a lower genetic liability in females (*i*.*e*., higher susceptibility to CP due to genetic variation). Several genome-wide association studies (GWAS) for CP are published; however, all are sex-combined cohorts (Leslie et al. 2016; Beaty et al. 2011; Butali et al. 2019; Huang et al. 2019; He et al. 2020; Robinson et al. 2023). Stratification by sex in CL/P has led to discovery of novel associated sex-specific factors (Carlson et al. 2018; Awotoye et al. 2022), but no such study has been conducted for CP. Although the observed sex bias is opposite for CP, it is plausible there are similar sex-specific risk factors. One genome-wide interaction study identified SNPs associated with environmental exposures such as maternal smoking, alcohol intake, or vitamin use (Beaty et al. 2011), but none have specifically examined gene-by-sex interactions.

Taken altogether, we hypothesized sex may influence the occurrence of CP. To test this hypothesis, we first performed sex-stratified GWAS, followed by gene-by-sex (GxS) interaction analysis on 435 cleft palate case-parent trios. After identification of 13 loci with significant interactions, we also tested the association of the predicted genetically regulated gene expression with OFCs in the full cohort and by sex for the nearest genes for these significant loci.

## Methods

### Study population and phenotyping

The study population was recruited from domestic and international sites in North America (United States, Puerto Rico), Europe (Spain, Turkey, Hungary), South America (Argentina), Asia (China, Singapore, Taiwan, the Philippines), and Africa (Nigeria, Ghana). In total, 435 case-parent trios were ascertained through probands affected with CP. We did not exclude subjects based on parent affection status or the presence of additional clinical features. There were 19 trios (4.4%) with an affected parent and 46 trios (10.6%) with minor or major clinical features in addition to CP. Proband sex was confirmed genetically for all cases during quality control processes.

### Sample preparation and whole genome sequencing

Whole genome sequencing was performed at the Center for Inherited Disease Research (CIDR) at Johns Hopkins University (Baltimore, MD). Prior to sequencing, a Fragment Analyzer system was used to test samples for adequate quantity and quality of genomic DNA, which were then processed with an Illumina InfiniumQCArray-24v1-0 array to confirm sex, relatedness, and known duplicates. For each sample, 500-750ng of genomic DNA was sheared to 400-600bp fragments, then processed with the Kapa Hyper Prep kit for End-Repair, A-Tailing, and Ligation of IDT (Integrated DNA Technologies) unique dual-indexed adapters according to the Kapa protocol to create a final PCR-free library.

Sequencing was performed on 150bp paired-end reads on a NovaSeq 6000 platform with base calling through the Illumina Real Time Analysis software (version 3.4.4). Files were demultiplexed from binary format (BCL) to individual fastq files with Illumina Isa s bcl2fastq (version 1.37.1) and aligned to the human hg38 reference sequence (https://www.ncbi.nlm.nih.gov/assembly/GCF_000001405.39). Alignment, variant calling, and quality control was done with the DRAGEN Germline v3.7.5 pipeline on the Illumina BaseSpace Sequence Hub platform, which produced single sample VCF files. Contamination for any cross-human sample was checked with the DRAGEN contamination detection tool. We performed genotype concordance with existing array-based genotypes using CIDRSeqSuite (version 7.5.0), and genotype concordance checks amongst replicate samples was performed in Picard GenotypeConcordance (Picard 2019). After data quality steps, samples with at least 80% of the genome at 20X coverage or autosomal coverage at 30X underwent joint calling to generate a multi-sample VCF.

### Quality control

Variants underwent quality control in the multi-sample VCF. Only variants aligning to the standard chromosomes (1-22, X, Y) were included. Variants with any flag other than “PASS” and a minor allele count (MAC) of <2 were removed. Genotypes were set to missing if the quality score was <20 or had a read depth of <10, and then variants with missingness >10% were removed. Sample-level metrics were then evaluated for heterozygous/homozygous ratio, transition/transversion (Ts/Tv) ratio, and silent/replacement rate. Outlier samples with values outside of 3 standard deviations from the cohort mean were discarded. Samples with high missing data (>5% missing) were removed.

### Statistical analysis

We performed GWAS using transmission disequilibrium tests (TDT) on the full cohort and stratified by proband sex. The TDT uses the rate of transmission of the minor allele to determine association. This is beneficial as it is not confounded by population stratification, however, it is only informative at sites for which a parent is heterozygous. TDTs were performed using the R package *trio* (Schwender H 2023) for trios in which all individuals had a genotype missing rate of <5% and did not exceed a Mendelian error rate of 2%. Variants were removed based on the following filters: minor allele frequency (MAF) <3%, Mendelian error rate >0.1%, Hardy-Weinberg disequilibrium as determined by exact test p-value of <1x10^-6^, and missingness rate of >5%. Relative risk is reported in respect to the alternate allele. We considered p<1x10^-5^ to be the threshold of suggestive significance and p<5x10^-8^ of genome-wide significance.

We then performed a TDT-based gene-by-sex interaction analysis in *trio*, using the colGxE() function, which uses a two-step procedure as described by Gauderman et. al (Gauderman et al. 2010). We first evaluated all variants using a likelihood ratio test with two degrees of freedom (LRT 2df), which simultaneously tests the SNP main effect and environmental interaction. For this function, a χ2-distribution is used as the asymptotic null distribution for p-values determination.

SNPs reaching suggestive levels of association (p<1x10^-5^) were then evaluated specifically for gene-by-sex interactions. The GxS relative risk (RR) is reported as the RR in females divided by the RR in males. Because these tests are considered independent of one another, we used the number of suggestive SNPs in the LRT 2df test to calculate the significance threshold for the GxS step based on a Bonferroni correction (0.05/71 SNPs; p<7.04x10^-4^). We performed a *post hoc* power calculation in Quanto (version 1.2.4) to evaluate our power to detect interactions in the same direction with different magnitude or antagonistic interactions.

### Genetically regulated gene expression imputation and association testing

For each test gene, we first trained sex-specific gene expression imputation models with cis-SNPs as predictors using GTEx V8 reference data (31) for all 49 tissues in female samples and 48 tissues in male samples. According to the GTEx eQTL analysis protocols, covariates including sequencing protocol (PCR-free), sequencing platform (NovaSeq 6000), top five genotype principal components, and top probabilistic estimation of expression residual (PEER) factors (32) were regressed out from the gene expression quantitative traits (log2 transformed transcripts per million). For each tissue, we used the TIGAR-V2 tool (33) to train both nonparametric Dirichlet process regression (DPR) model and penalized the linear regression model with an Elastic-net penalty. Second, the trained sex-specific gene expression imputation models, i.e., the effect sizes of cis-SNP predictors (referred to as eQTL weights), were used to impute genetically regulated gene expression using the corresponding sex-specific GWAS data. For each gene, the sex-specific association between the genetically regulated gene expression (imputed by the DPR or by the Elastic-Net model) and CP were tested by the S-PrediXcan Z-score statistic (34) as implemented in the TIGAR-V2 tool. Third, for each gene, sex-specific test p-values based on the DPR and the Elastic-Net model were then combined by using the aggregated Cauchy association test (ACAT) (35). Previous studies have shown the ACAT method can improve power by leveraging multiple tests based on different statistical models of the same null hypothesis. As a result, we obtained sex-specific ACAT p-values for testing the association between the genetically regulated gene expression of test genes and the CP phenotype. A significant ACAT p-value means the genetic effect of the test gene (aggregated over multiple test cis-SNPs with non-zero eQTL weights) on CP is potentially mediated through the corresponding gene expression.

### Rare Variants

A bed file for hg38 coordinates for *LTBP1* was generated using the UCSC Genome Browser. All variants within this region were extracted from a multisample VCF with vcftools (version 0.1.13) followed by annotation with Annovar (version 201910) (Wang, Li, and Hakonarson 2010). The commands “dbnsfp42a” and “revel” were included to obtain SIFT, CADD, and REVEL scores for all variants. Additionally, prediction values from AlphaMissense (Cheng et al. 2023) were accessed from the publicly available data. Predicted damaging thresholds were as follows: SIFT ≤0.05, CADD ≥20, REVEL ≥0.75, and AlphaMissense ≥0.564.

## Results

Using a dataset of 435 trios, we performed sex-stratified TDTs of 262 female and 173 male probands based on genetic sex (Fig 1, Online Resource 1, 2). For all evaluations, we tested 6,774,204 SNPs. There were 16 SNPs in 9 loci of suggestive significance in the female analysis, and 3 SNPs in 3 loci in males (Table 1). The significant loci from each test were non-overlapping between sexes—that is, they were only significant in either male or female probands. Additionally, each of these loci were more significant with higher odds ratios (ORs) within the sex-stratified TDTs than compared to the full cohort (Online Resource 3). To explore these sex-specific differences more formally, we next performed a GxS analysis to find sex-specific interactions.

**Table 1:**
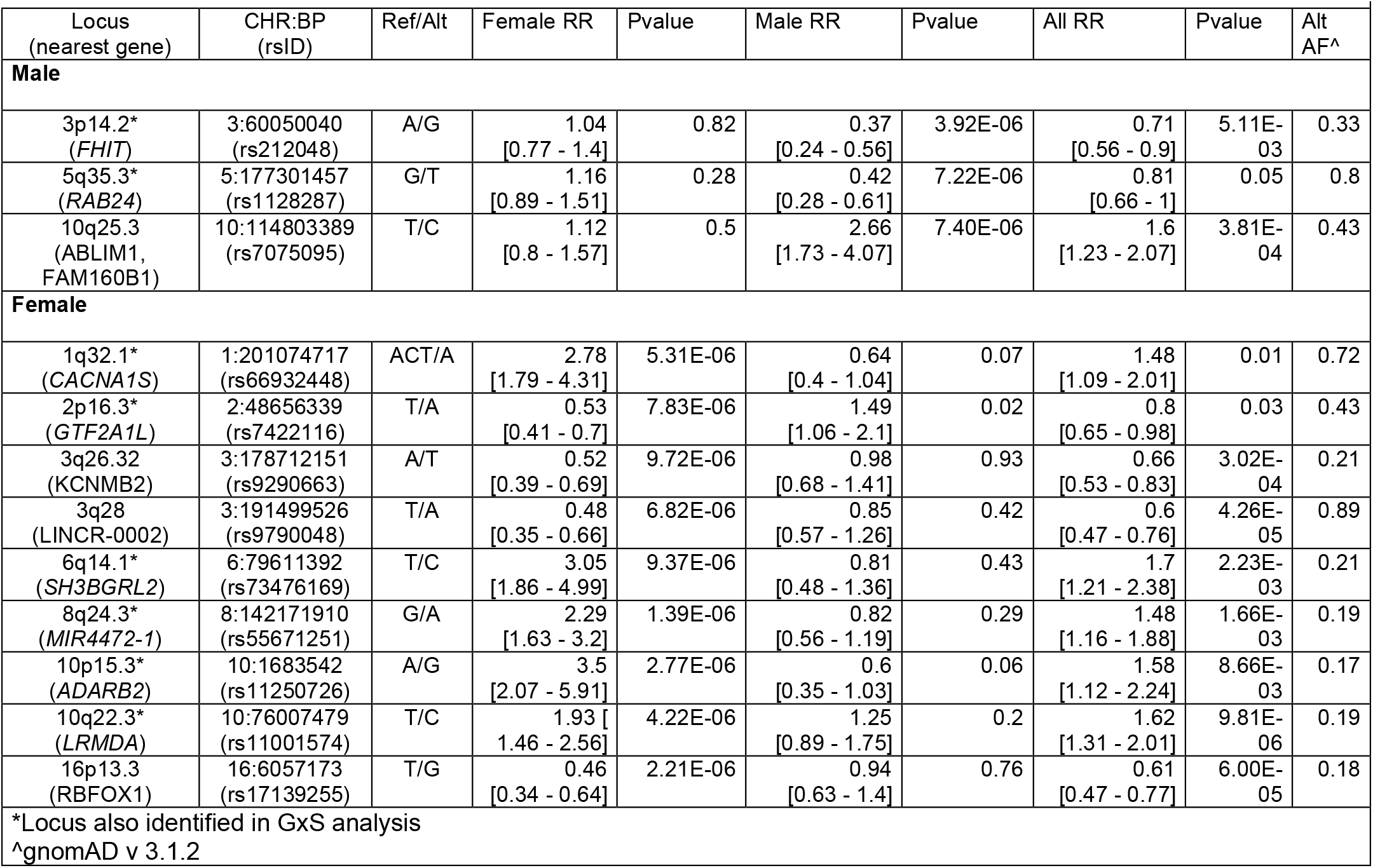
Loci of suggestive significance from sex stratified TDTs.

**Fig 1.**
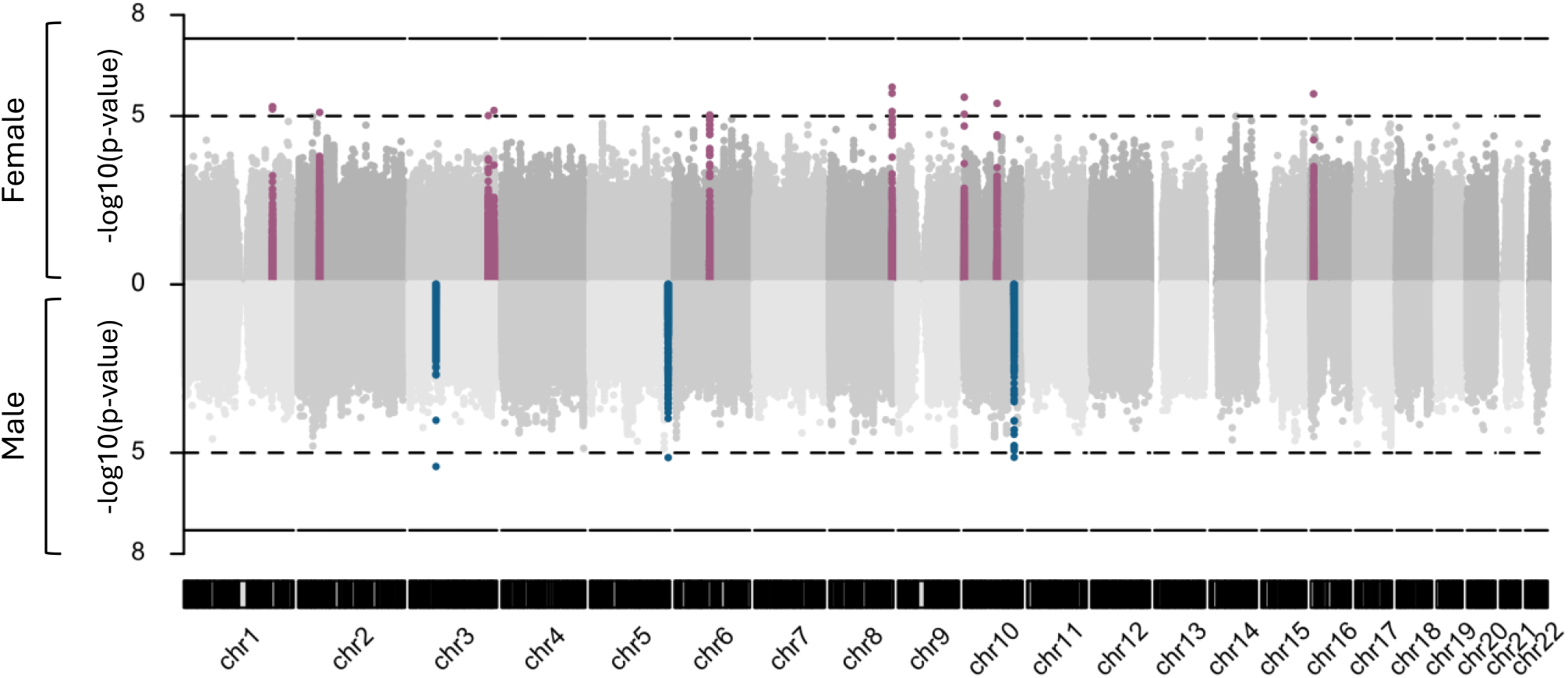
Miami plot highlighting differences in suggestive SNPs in females (top) and males (bottom). The dotted line represents suggestive significance at p=1x10^-5^, and the solid line represents p=5x10^-8^

We first tested SNPs with the LRT 2df model of association considering both gene and GxS effects (Online Resource 4). There were no SNPs at genome-wide significance in this step, though there were 71 SNPs in 32 regions with p<1 x 10^-5^ (suggestive significance threshold) (Table 2). Of those, 26 SNPs in 13 regions were considered significant for gene-by-sex interactions (Bonferroni correction for 71 SNPs, p<7.04 x 10^-4^) (Online Resource 5). We noted that for all GxS significant SNPs, the direction of effect was opposite in males versus females (Fig 2a, b). A *post-hoc* power analysis shows given our sample size, we have good power to detect large differences (including opposite effects) when SNPs have a MAF of at least ∼15%. However, we have very low power to detect modest interactions, or even large differences at lower MAFs (Online Resource 6). We did not detect significant GxS interactions in the remaining 45 SNPs in 19 regions, suggesting that these associations are primarily driven by the effect of the genotype rather than sex-specific interactions (Fig 2c).

**Table 2:**
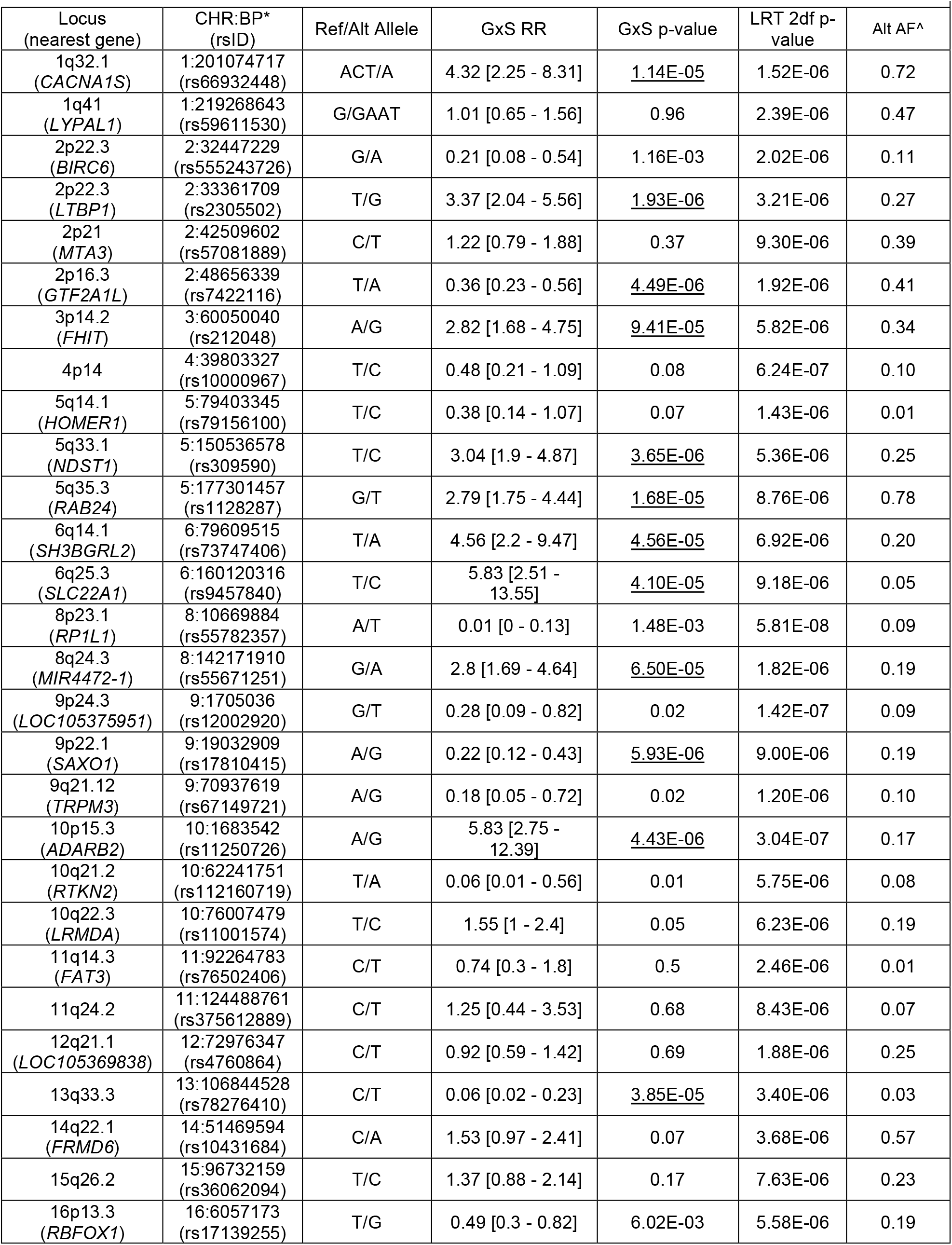
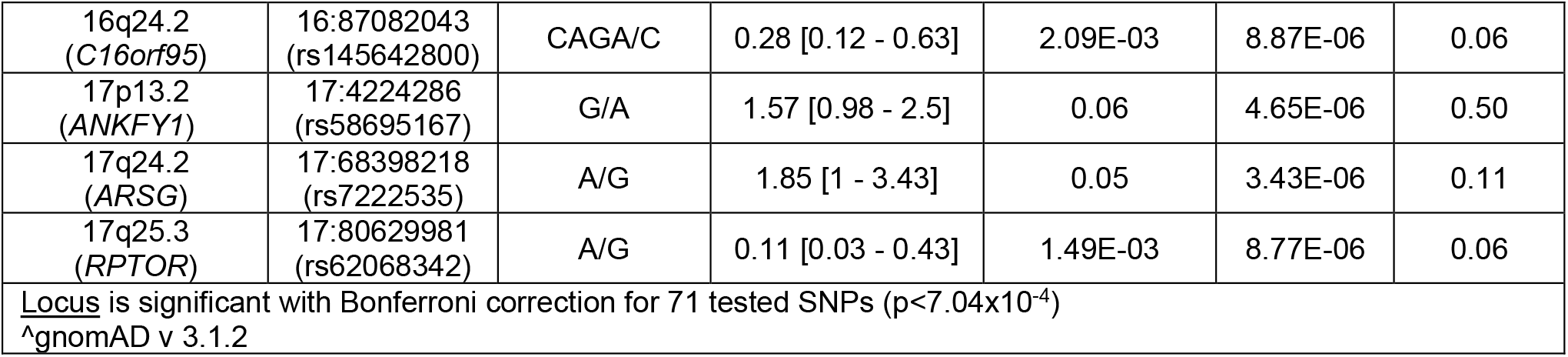
Loci of suggestive significance in the LRT 2df.

**Fig 2.**
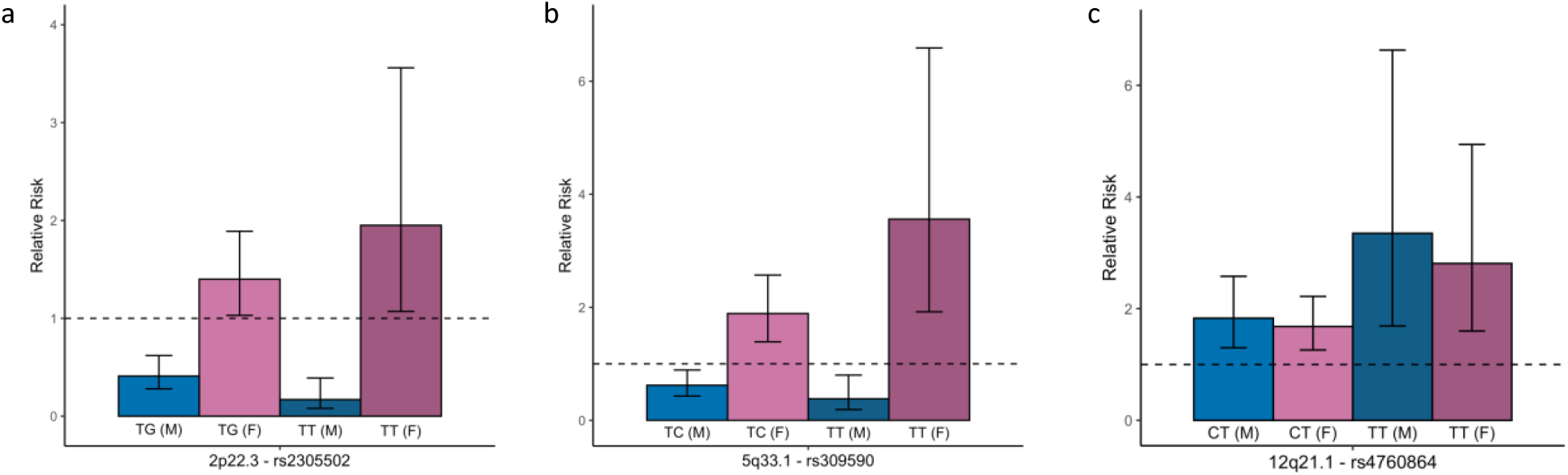
Relative risks for males and females demonstrating GxS effect SNPs at (A) 2p22.3 and (B) 15q21.1, and (C) gene-driven SNP at 112q21.1 with 95% confidence intervals. The dotted line at 1 represents null risk. Light blue and dark blue represent heterozygous and homozygous males, respectively while light and dark pink represent heterozygous and homozygous females, respectively

For the 13 loci from the sex-stratified TDTs (Table 1), two of the three loci from the male-specific analysis, and six of nine loci from the female-specific analysis, we found significant for GxS effects. Using the results from all four tests, we compared the relative risks for the ten most significant SNPs in the GxS to those from the sex-stratified and sex-combined TDTs, shown in Fig 3.

**Fig 3.**
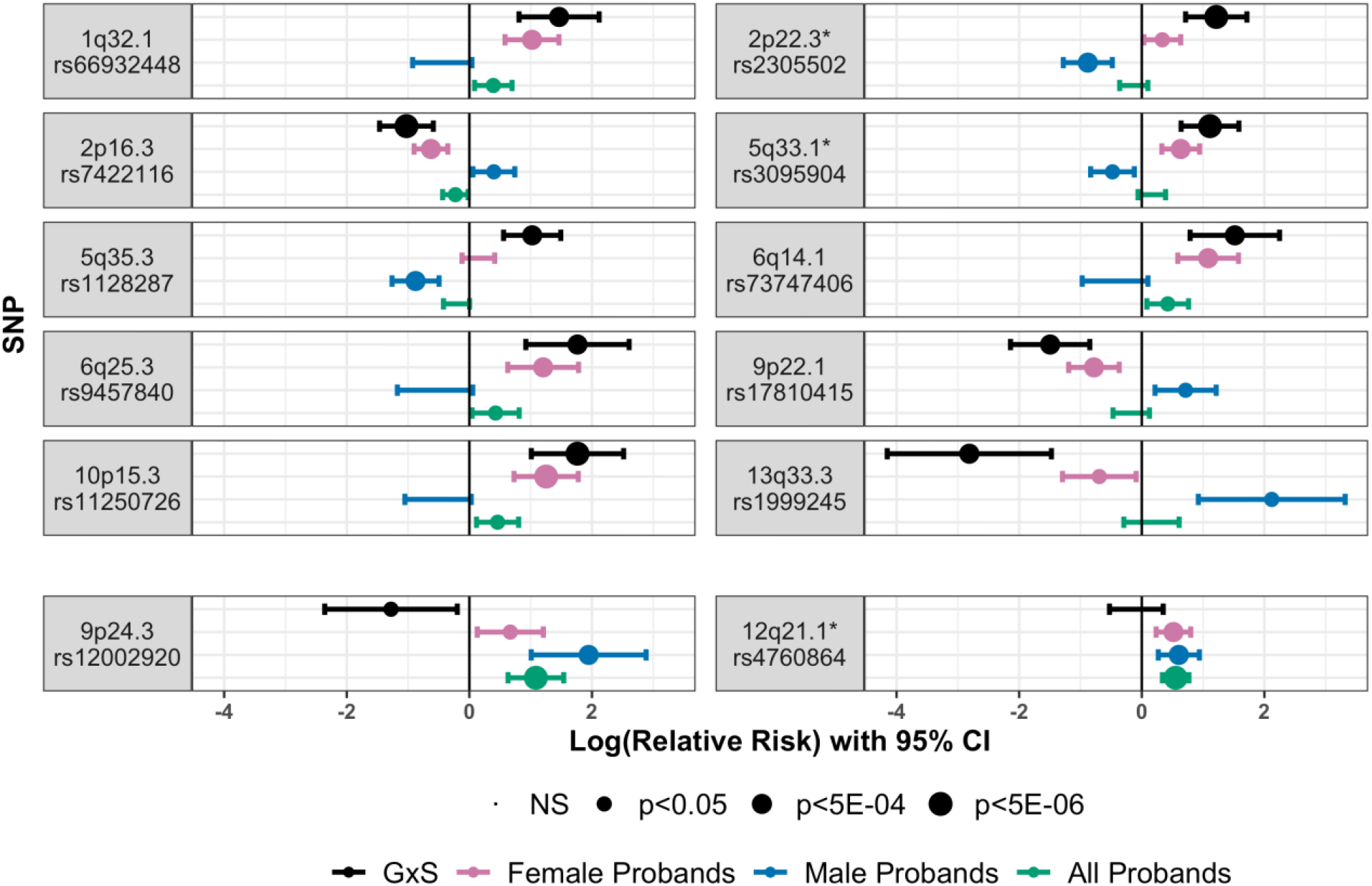
Comparing relative risks with 95% confidence intervals across the top 10 SNPs for GxS effects (top) and two gene-driven SNPs (bottom) to relative risk from TDTs in the full cohort and stratified by sex. Risks are shown for GxS (black), female only TDT (pink), male only TDT (blue), and full cohort TDT (green). An asterisk next to the locus name denotes relative risk by heterozygosity shown in Fig 2

The most significant finding for GxS interaction was at 2p22.3, within intron 28/33 of latent TGF-beta binding protein 1 (*LTBP1*). We expanded our analysis of *LTBP1* to look at rare variants (MAF <0.05%) in the full cohort and identified 21 unique protein-altering variants in 25 probands (Online Resource 7). 14 variants in 15 probands had at least one damaging prediction as determined by CADD, REVEL, or AlphaMissense scores. We performed a Fisher’s exact test on the difference in the number of predicted damaging rare variants in male versus female probands and found these were not statistically significant (p=0.22).

Lastly, there is evidence for differential expression of *LTBP1* between sexes in both the pituitary of mice (Recouvreux et al. 2013) and whole blood of humans (Recouvreux et al. 2013). Therefore, we evaluated the association between the sex-specifically imputed genetically regulated gene expression and the CP phenotype for 12 genes present at the GxS significant loci. We also evaluated the association between the imputed genetically regulated gene expression and the CP phenotype in all samples. There were 11 genes with nominal significance (p<0.05) in at least one of the tested tissues (Table 3). After correction for 12 loci, there were only two loci remaining significant: *GTF2A1L* for females in the substantia nigra of the brain (p=0.0010), and *LTBP1* for females in cultured fibroblast cells (p=0.0013). The latter association was also observed to a lesser extent for the full cohort (p=0.0125).

**Table 3:**
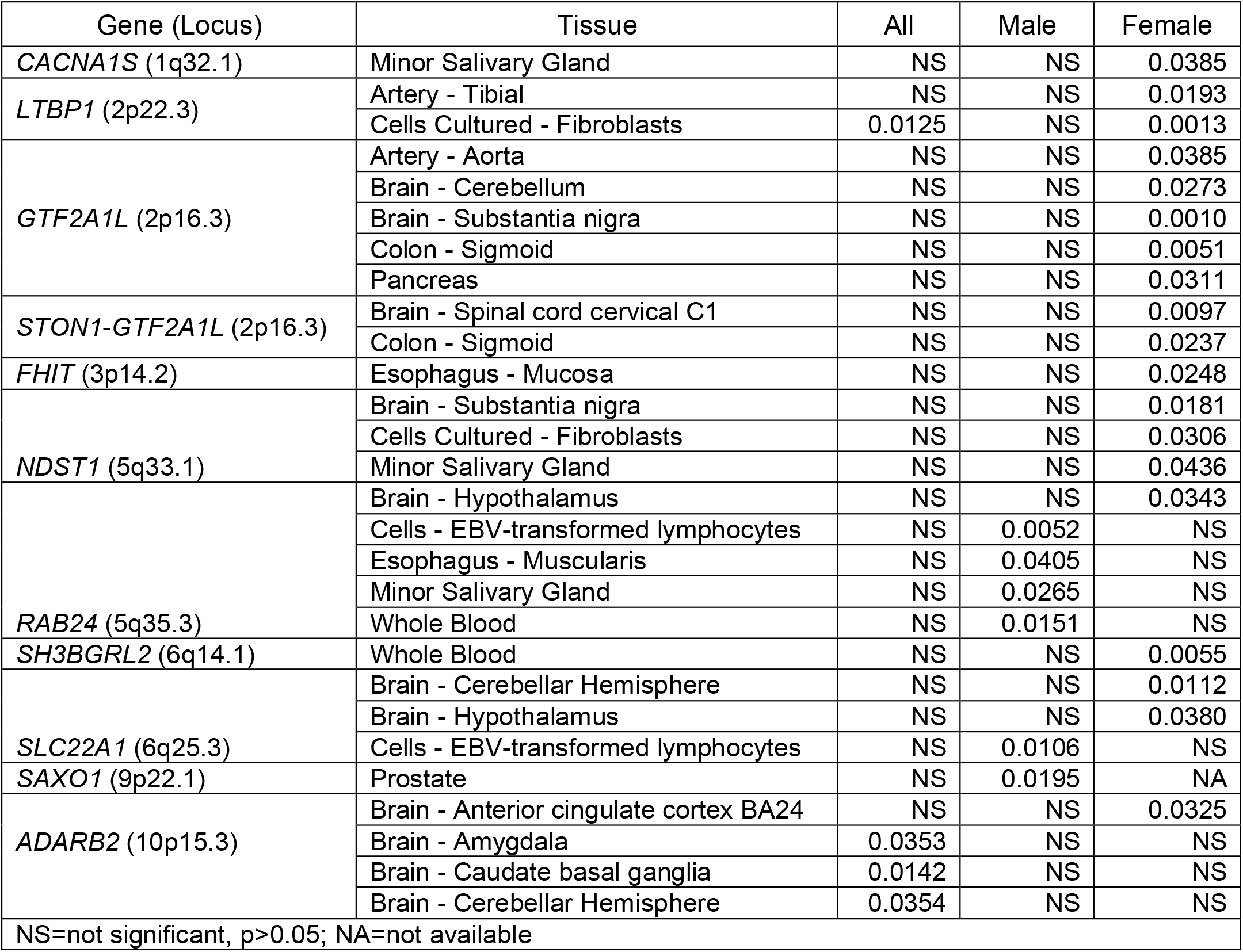
Tissues and genes with significant association between imputed gene expression and phenotype.

## Discussion

CP is more prevalent in females than males, yet little is known about what contributes to these sex-specific risks. Here, we initially found 12 loci with suggestive evidence of association with CP when stratifying our cohort by proband sex, followed by identification of 13 loci with suggestive evidence of gene-by-sex interactions in CP probands. In all instances, the identified SNPs increased risk of CP for one sex while appearing to reduce the risk for the other. Given our sample size, this was expected as we have more power to detect these large differences in effects; much larger sample sizes would be needed to detect effects of different magnitude in the same direction in males and females. We also found that most loci were associated with a higher relative risk in females compared to males, which supports the epidemiological data where females are more commonly affected.

In the LRT 2df analysis, we identified 13 loci associated with CP but not significant for GxS interactions. This suggests that these associations are primarily gene-driven, where the genotype influences the risk for CP regardless of patient sex. We reported the effects of these gene-driven SNPs in our previously published GWAS results for the full cohort (Robinson et al. 2023), and our results were consistent across both studies, as expected. However, none of the loci with significant GxS interactions were significant in that study, as the risks in males and females likely negated each other. We also compared these loci to sex-specific risks previously reported for OFCs (Carlson et al. 2018; Awotoye et al. 2022), but none were replicated in our cohort. Altogether, these findings illustrate heterogeneity in genetic architecture both between CP and CL/P as well as between males and females, and highlight the importance of investigating sex -specific differences and/or GxS interactions in CP. While our study contributes to the knowledge of the genetic basis of CP and sex-specific risks, it is not without limitation. Our sample size of 435 trios is underpowered to identify gene-by-sex interactions in the same direction but with differences in the magnitude of risk. Further, due to the use of TDTs for case-parent trios, we are restricted to evaluation of sites for which parents are heterozygous.

Our top finding was an intronic SNP in *LTBP1* at 2p22.3. There are several lines of evidence suggesting that *LTBP1* could play a role in palatogenesis. LTBP1 is an extracellular matrix protein and a component of the large latent complex (Peeters et al. 2022). This complex binds to a second complex of latency associated proteins and mature TGF-B, working to regulate its biological availability in the extracellular matrix. Disruption of the TGF-B signaling pathway is associated with CP (Iwata, Parada, and Chai 2011; Taya, O’Kane, and Ferguson 1999), so dysregulation caused by variants in *LTBP1* could hypothetically lead to disrupted palatogenesis. We further identified 21 unique, rare variants (MAF <0.5%) in 12 males and 13 females. Of the 21 total, 15 were predicted to be damaging by at least one *in silico* tool. These were found in 9 males and 6 females, although this difference was not significant with a Fisher’s exact test.

From a clinical standpoint, bi-allelic truncating variants in *LTBP1* have been reported as causal for autosomal recessive cutis laxa syndrome type IIE (OMIM: 619451). In addition to connective tissue abnormalities, affected individuals present with craniofacial dysmorphisms including high arched palates in 62.5% of cases (Pottie et al. 2021). Further, LTBP1 directly competes with GARP (glycoprotein A repetitions predominant, encoded by *LRRC32)*, a type I membrane protein that plays a role in maturation, processing, and tethering of TGF-B to the cell surface (Tran et al. 2009). Both LTBP1 and GARP compete for binding via a disulfide link to the same cysteine (Cys-4) on proTGFB1, and they direct TGF-B to either the cell surface or deposited within the ECM (Wang et al. 2012). Recently, homozygous variants in *LRRC32* have been reported to cause a cleft palate syndrome (Hexner-Erlichman et al. 2022), suggesting imbalance in this process can disrupt palatogenesis.

In support of clinical data, morpholino knockdown of *ltbp1* in zebrafish causes severe jaw malformations in which the mandible has a reduced length and width. Similarly, knockout of *Ltbp-1* in mice resulted in a modified facial profile including a shortened maxilla and mandible, as well as reduced cellular activation and differentiation of myofibroblasts, supporting reduced availability of *Tgfb* when *Ltbp-1* is disrupted (Drews et al. 2008). In humans, shortened mandibles are associated with CP in Pierre Robin sequence (Resnick et al. 2018). Our cohort included 18 trios reported to have Pierre Robin sequence and an additional 31 with clinical features consistent with syndromic CP. While this was too few to perform a separate analysis, we carried out sensitivity analyses in which we repeated the GxS for *LTBP1* without PRS cases, as well as without PRS cases or any suspected to be syndromic. Our results did not meaningfully change, suggesting that—in this study—Pierre Robin or syndromic cases are not driving the signal in this gene.

Although the mechanism by which variants in or around *LTBP1* mediate their effect on a sex-specific basis is unknown, there is evidence for sex-based differences in expression of this gene. A study in mice found that *Ltbp1* expression in the pituitary was lower in females than males, and that both sexes showed reduced expression in response to estradiol, suggesting a role for sex hormones in this differentiation (Recouvreux et al. 2013). A similar finding has been shown in human peripheral blood, with *LTBP1* expression belonging to the top 1% of sex-biased genes (based on log fold change) where males had higher levels of LTBP1 mRNA than females (Jansen et al. 2014). To explore this based on our GWAS data, we performed an association analysis for CP and the imputed genetically regulated gene expression and found a female-specific significant association for *LTBP1* in cultured fibroblast cells. Despite a lack of access to a more relevant tissue (i.e., palatal tissue), the association of *LTBP1* expression with CP in female probands is of interest as fibroblasts have many of the same traits as mesenchymal stems cells, including an ability to differentiate into chondrocyte and osteoblasts (Denu et al. 2016).

In summary, we have demonstrated that there are gene-by-sex interactions for CP which cannot be detected in combined-sex cohorts as opposite allelic risks can negate each other. We also identified *LTBP1* as a gene of interest for CP risk, particularly for females, and show that it may mediate its effect on the phenotype via gene expression. In conclusion, sex-specific effects contribute to the genetic heterogeneity of CP and should be further explored as sample sizes continue to grow.

## Supporting information

Supplemental Resources 1-6

Supplemental Resource 7

## Data Availability

Sequence and phenotype data is available from the Database of Genotypes and Phenotypes (dbGaP) under study accession phs002220.v1.p1.

## Acknowledgements

We would like to thank participants and their families, without whom this research would not be possible. We would also like to thank Elena Serna and Rosa Martinez for collecting samples and maintaining the UTHealth database. Sequencing services were provided by the Center for Inherited Disease Research (CIDR). CIDR is fully funded through a federal contract from the National Institutes of Health to The Johns Hopkins University, contract number HHSN268201700006I. Patient recruitment, assembly of phenotypic information, sequencing services, and data analysis were supported by National Institutes of Health (NIH) grants: X01-HG010835 (EL), R01-DE016148 (MM, SW), R01-DE030342 (EL), R01-DE011931 (JH), R01-DE028300 (AB), R01-DE014581 (TB), R37-DE008559 (JM), R00-DE024571 (CB), S21-MD001830 (CB) U54-GM133807 (CB), T32-GM008490 (KR), F31-DE032588 (KR). This work was in part supported through cooperative agreements under PA 96043 from the Centers for Disease Control and Prevention to the Centers for Birth Defects Research and Prevention participating in the National Birth Defect Prevention Study. The findings and conclusions in this report are those of the authors and do not necessarily represent the official position of the Centers for Disease Control and Prevention or the California Department of Public Health.

## Competing Interests

The authors have no relevant financial or non-financial interests to disclose.

## Ethics Approval

This study was performed in line with the principles of the Declaration of Helsinki. Institutional review board approval was obtained for each local recruitment site and coordinating center: University of Iowa, University of Pittsburgh, and Emory University.Informed consent was obtained from either the parents or all individual participants included in the study.

## Notes

### Competing Interest Statement

The authors have declared no competing interest.

### Author Declarations

Institutional review boards of University of Iowa, University of Pittsburgh, and Emory University gave ethical approval for this work.

