## Supplemental Resources 1-6 for "Genome-wide study of gene-by-sex interactions identifies risks for cleft palate"

Online Resources

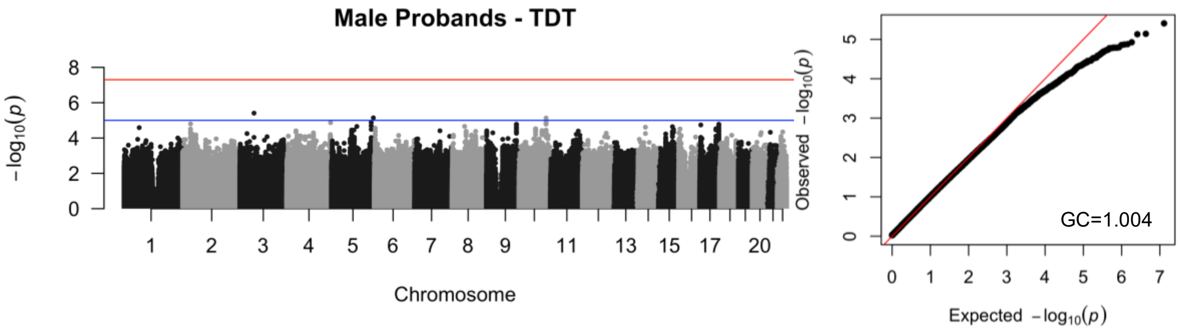


Online Resource 1: Manhattan plot for male probands (left) with qqplot of p-values (right). The red line represented genome-wide significance at p=5E-08, and the blue line represents suggestive significance at p=1E-05.

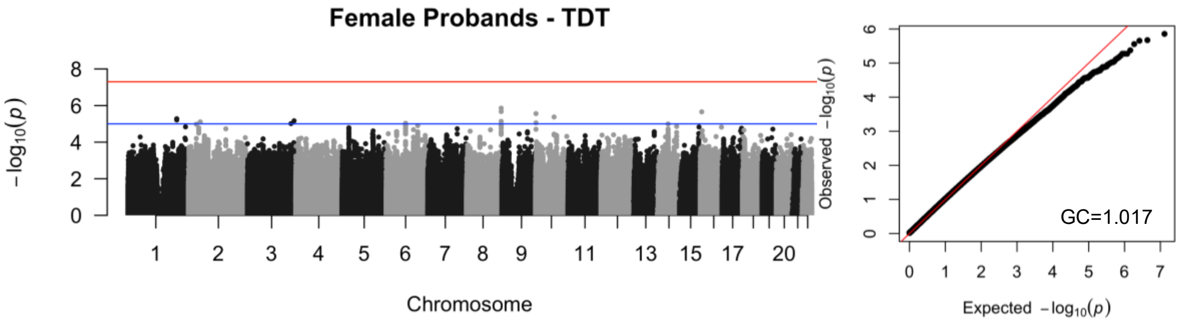


Online Resource 2: Manhattan plot for female probands (left) with qqplot of p-values (right). The red line represented genome-wide significance at p=5E-08, and the blue line represents suggestive significance at p=1E-05.

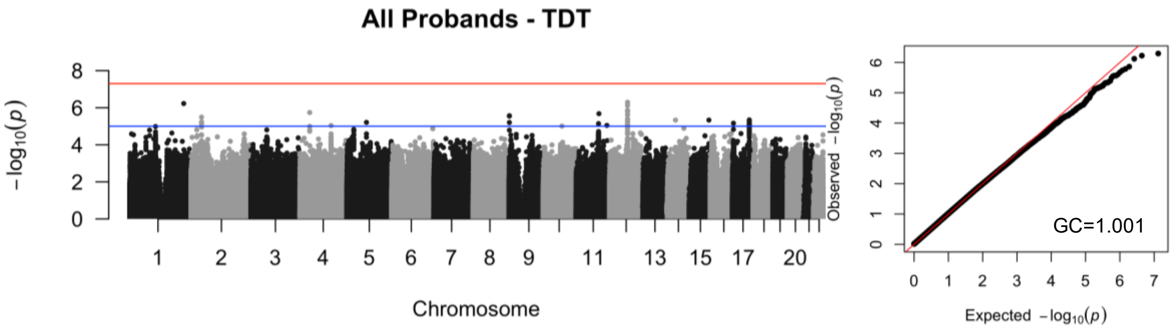


Online Resource 3: Manhattan plot for all probands (left) with qqplot of p-values (right). The red line represented genome-wide significance at p=5E-08, and the blue line represents suggestive significance at p=1E-05.

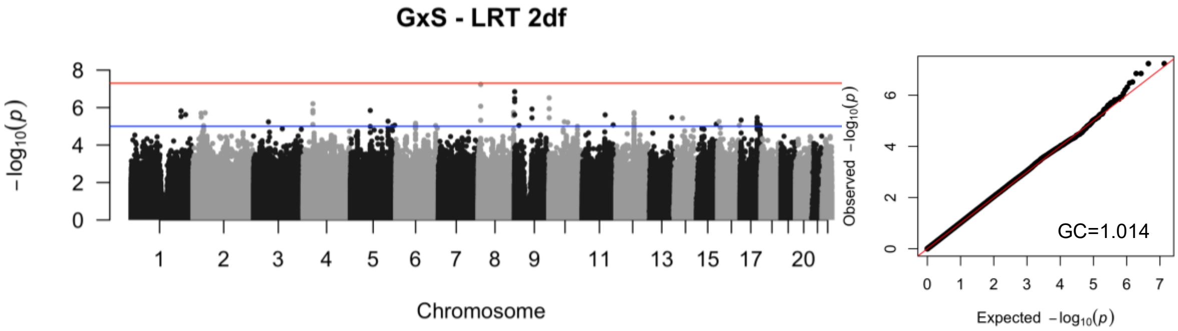


Online Resource 4: Manhattan plot for the LRT2df (left) with qqplot of p-values (right). The red line represented genome-wide significance at p=5E-08, and the blue line represents suggestive significance at p=1E-05.

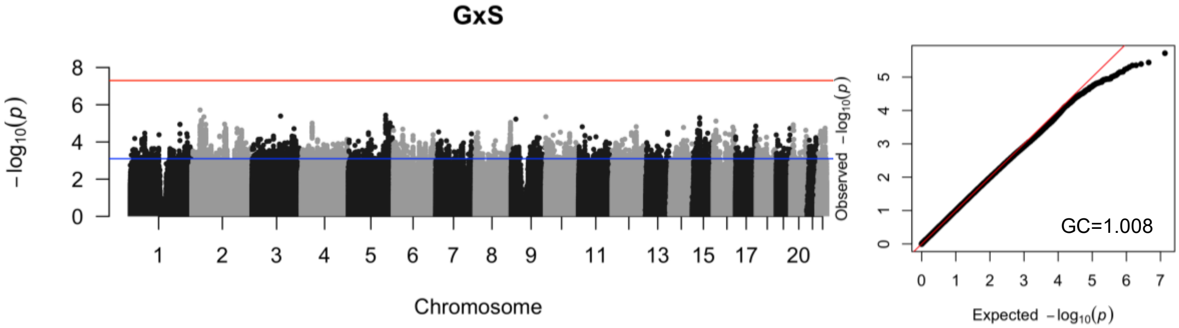


Online Resource 5: Manhattan plot for GxS interactions (left) with qqplot of p-values (right). The red line represented genome-wide significance at p=5E-08, and the blue line represents suggestive significance at p=1E-05.

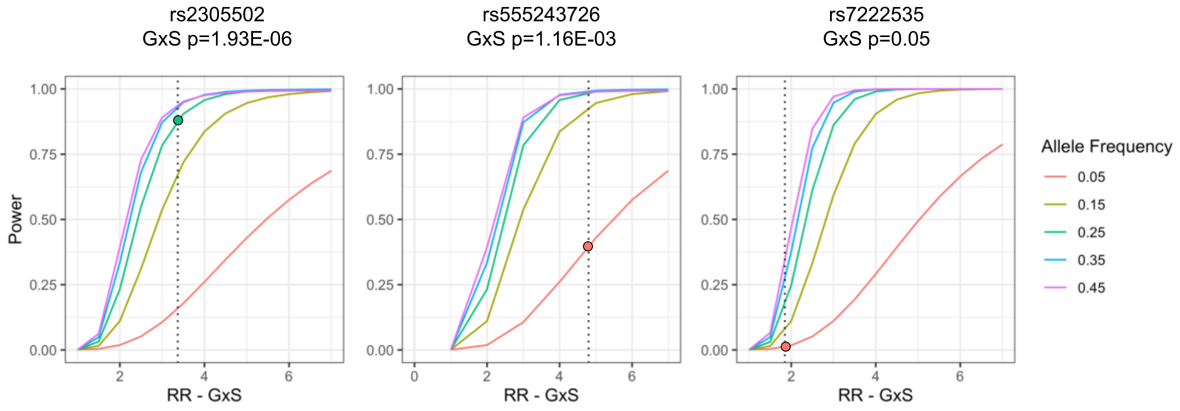


| Parameters | Parameters | Parameters |
|---|---|---|
| $R_G$ = 0.4 $R_E$ = 1.4 | $R_G$ = 1.05 $R_E$ = 5.0 | $R_G$ = 0.35 $R_E$ = 0.65 |
| $R_{GE}$: 1.0-7.0 | $R_{GE}$: 1.0-7.0 | $R_{GE}$: 1.0-7.0 |

Online Resource 6: *Post hoc* power calculations for the most significant SNP for GxS effect (left), the SNP just below the significance threshold for GxS effect (middle), and a SNP with no significant effect for GxS (right). In each plot, the dotted vertical line represents the output $R_{GE}$ determined by Trio, and the colored dot represents each SNPs respective MAF at the intersection of its $R_{GE}$ along the line of power of detection. All SNPs were tested using a population risk of 0.00058 and ‘environmental exposure’ of 0.5, with additional SNP-specific corresponding input parameters used for each SNP are listed below the image.
